# Managing Low Back Pain in Rural Uganda: A Qualitative Study Exploring the Perspectives and Practices of Frontline Health Workers regarding LBP Management in Primary Care

**DOI:** 10.1101/2024.05.15.24307404

**Authors:** Paul Harscouët, Gloria Ndekezi Chimpaye, Herman Kazibwe, Jerome Kabakeynga, Denise O’Callaghan, Catherine Blake, Brian Caulfield, Cliona O’Sullivan

## Abstract

**BACKGROUND AND AIMS:** Low-back pain (LBP) is the main cause of years lived with disabilities (YLDs) worldwide and the second cause of YLDs in Uganda. In 2019, it was responsible for 7.4% of global YLDs and 5% of YLDs in Uganda. LBP takes a significant toll on people’s quality of life and disproportionately affects lower socioeconomic classes, elders, and women. In rural Uganda, LBP is managed in health centres by clinical officers and nurses with limited resources. This study aims to understand the perspectives and practices of these health workers.

**Method:** A qualitative design using semi-structured focus-group discussions was employed. Purposive sampling allowed us to identify relevant participants based on their roles as healthcare professionals working in primary care context in rural South-West Uganda. Data was analysed using thematic analysis.

**Findings:** LBP is a common and persistent complaint among patients presenting to rural health centres in Uganda. Manual labour and female specific health conditions were deemed to be common causes. There was a strong reliance on medication prescription, coupled with X-ray diagnosis, with little emphasis on education or exercise. Finally, findings highlighted major barriers for patients within the referral system to hospital care or rehabilitation.

**Discussion:** Education and training of frontline clinicians in terms of appropriate prescribing and rehabilitation for LBP is crucial. Evidence-based rehabilitation interventions need to be developed and adapted so that they can be delivered within the time and resource constraints of the health workforce, ensuring that they are acceptable and effective to patients in the context of rural Uganda.

## Introduction

Low-back pain (LBP) is the main cause of years lived with disabilities (YLDs) worldwide and the second cause of YLDs in Uganda [1–2]. It affected an estimated 619 million persons globally in 2020 and in 2020, it was responsible for 7.7% of global YLDs and 5% of YLDs in Uganda [2–3]. It is a debilitating condition with a lifetime prevalence of 84% and a chronic form prevalence of 23% [4]. The prevalence of LBP is higher in high-income countries but the demographic projections of low- and middle-income countries, (LMICs) such as increased life expectancy and ageing, will be consequential to the rise of LBP prevalence [2, 4]. In Uganda, the prevalence of back pain among older persons is estimated to be 64.7% with women being significantly more affected than men [5]. Low-back pain has a socio-economic gradient, with lower socioeconomic status being associated with a higher prevalence [6, 7].

Chronic LBP takes a significant toll on people, affecting “various domains of daily life from basic self-care activities to advanced and complex social interactions, work and leisure activities and eventually leading to poor quality of life” [6]. A review of qualitative studies investigating the impact of LBP on people’s lives described the consequences of LBP as “extending beyond functional considerations, and pervading many aspects of patients’ lives” [8]. They summarized the impact of LBP under five main themes: activities, relationships, work, stigma, and changing outlook [8]. However, most of these studies were conducted in high-income countries (40/42). Older persons tend to have greater disability resulting from LBP than younger people [9]. In urban Uganda, LBP is associated with a loss of working days and a reduction in the overall quality of life [10]. Despite the growing burden of musculoskeletal impairment, musculoskeletal health tends to be under-prioritised in LMICs’ non-communicable health policies [11].

LBP usually has no known pathoanatomical cause, often referred to as non-specific low back pain [9], however it can be the symptom of a severe pathology, such as spinal fracture, infection, malignancy, cauda equina syndrome, or radiculopathy [9, 12]. A study conducted in an orthopaedic clinic in urban Uganda among patients seeking LBP care as chief complaint found that 19.1% had nerve root compression and 17.2% of patients had a serious spinal pathology [13]. The frequency of serious spinal pathology in this study was higher than the 1% to 10% frequency reported in high-income contexts [9].

In Uganda, firstline care for patients with LBP in rural areas is delivered in health centres III and IV by clinical officers or nurses. Health centres III are found at the sub-county level and cover a population pool of 20 000 people. Health centres IV are found at the county level and cover a population pool of 100 000 people. If a serious pathology is detected, patients are then either managed locally for certain health conditions (e.g. brucellosis, extra-pulmonary TB) or referred to a hospital if more specialist care and investigation is warranted (e.g. risk of cancer). LBP management is not specifically addressed in the Uganda Clinical Guidelines where the only reference to LBP as a condition can be found under the domain of palliative care [14].

Rehabilitation, in the form of advice, education and exercise, is considered firstline care for non-specific LBP [15]. In Uganda, there is a severe rehabilitation workforce scarcity [16]. This scarcity is most important in rural areas where rehabilitation services are practically non-existent and the referral system faces many challenges, making these services inaccessible to rural populations. As a human right, rehabilitation is particularly important for people with disabilities, including those with LBP.

This study was undertaken as part of a larger project, called the BACKTRACK project, which aimed to develop technology-enabled care pathways, as a solution to improve LBP management in rural communities in Uganda. Specifically, this study sought to understand the perceptions and practices relating to LBP of primary care clinicians working in rural areas. It follows the Human Rights Council call to “increase rehabilitation-related research, especially in priority areas identified by WHO [World Health Organization], such as the types and impacts of different service delivery models, […] and facilitators and barriers to accessing rehabilitation” [17].

## Methods

### Context and approach

The BACKTRACK research project is co-led by researchers from Mbarara University of Science and Technology (MUST), Uganda and University College Dublin (UCD), Ireland. Our methodology reporting follows the COREQ guidelines for qualitative studies report [18].

### Study design

A phenomenological approach using qualitative design in the form of focus-group discussion was used to explore the practice of low-back pain management by healthcare workers in rural South West Uganda. Focus group discussion was deemed an appropriate method to facilitate deeper discussion, shared understandings and diverse viewpoints from clinicians working in different health centres, and enabled the research team to collect a broad range of views within a short time-frame.

### Participant recruitment

Ten health centres (levels III and IV) were randomly selected from a list of health centres that refer their patients to Mbarara Regional Referral Hospital (MRRH). Permission to recruit participants was granted by the relevant district health officers for the participating health centres.

Purposive sampling allowed us to identify relevant participants based on their roles as healthcare professionals working in primary care context in rural South-West Uganda, ensuring where possible there was a nurse and clinical officers was included from each participating health centre. All participants received an invitation to participate from MUST. Participants came from three different districts: Mbarara, Rwampara and Isingiro and worked at HC III and IV level. Every person contacted agreed to participate.

### Focus Groups

Three FGDs were conducted and facilitated separately by three researchers (COS, GNC, PH) in Mbarara in October 2022. The question set used to guide the FGDs was developed in advance, during several meetings with researchers and clinicians from the two universities involved in the BACKTRACK project and guided by the overall study aim. Prior to the focus groups, participants had read the participant information leaflet and signed a consent form. At the beginning of the FGD, the facilitators introduced themselves and reiterated the aim of the BACKTRACK project and the purpose of the focus groups. The focus group discussion was semi-structured, with the question set used as a prompt to initiate and guide the discussion. Participants were encouraged to discuss freely and openly. All FGDs were conducted in English, audio-recorded and lasted sixty to seventy-five minutes. No field notes were made. Transcripts were not returned to participants for comment nor correction.

### Data Analysis

We conducted a thematic analysis of the transcripts [19–21]. The data generated provided a rich description of perceptions and experiences, which facilitated the use of an inductive and constructivist approach at a latent level to facilitate identification of themes. Providing a rich description means that the “themes identified and developed here are an accurate reflection of the data set” instead of focusing on a particular aspect of it [19]. The inductive approach implies that the themes identified are strongly linked to the data itself, instead of being theory-driven. Analysing at the latent level goes beyond the semantic level to “ identify or examine the underlying ideas, assumptions, and conceptualizations -and ideologies-that are theorised as shaping or informing the semantic content of the data” [19]. The positionality of the three researchers, who conducted the FGDs and the data analysis, as physiotherapists with experience of managing LBP, (GN, PH, COS) and with a deep understanding of the Ugandan health system and context (GN) allowed analysis at a latent level.

Finally, a constructivist approach considers that “meaning and experience are socially produced and reproduced, rather than inheriting within individuals”. Three researchers, (GN, PH, COS) read and re-read the transcripts of the focus groups that they had facilitated (familiarisation, Step 1). Each researcher undertook line by line coding of their own focus-group discussion transcripts, (Step 2). The codes generated and corresponding quotes were recorded manually on Microsoft Excel and preliminary categories of data were identified by each researcher, (generation of themes, Step 3). Then, during two consensus meetings, preliminary themes were reviewed (Step 4) and agreement was reached on the meaning and naming of themes (Step 5). During the final phase, (write-up), codes and quotations were revisited to extract quotations to highlight the meaning of themes.

### Ethical approval

This study received ethical approval from the Mbarara University of Science and Technology Research Ethics Committee (MUST-2022-540) and from the Human Research Ethics Committee at University College Dublin (LS-LR-22-227-OSullivan).

## Results

The demographics of the participants can be found in table 1 below.

**Table 1–.** Participant’s Demographic Characteristics.

|  |  |  |
| --- | --- | --- |
| **Occupation** | Clinical Officer (n=8) | Registered Nurse (n=11) |
| **Gender** | Female (n=13) | Male (n=6) |
| **Health Centre (HC)** | Health Centre 4 (n=8) | Health Centre 3 (n=11) |
| **Age (years)** | $\mu=35.5$ ($\sigma=6.4$) | Min=29; Max=50 |

Five major themes were identified, some with sub-themes that interconnected and overlapped with other themes (Fig 1). The major themes were: (1) The burden of LBP, (2) Common aetiologies and causes, (3) Management of LBP, (4) Patient expectations, and (5) Challenges with referral pathways.

**Figure 1-.**
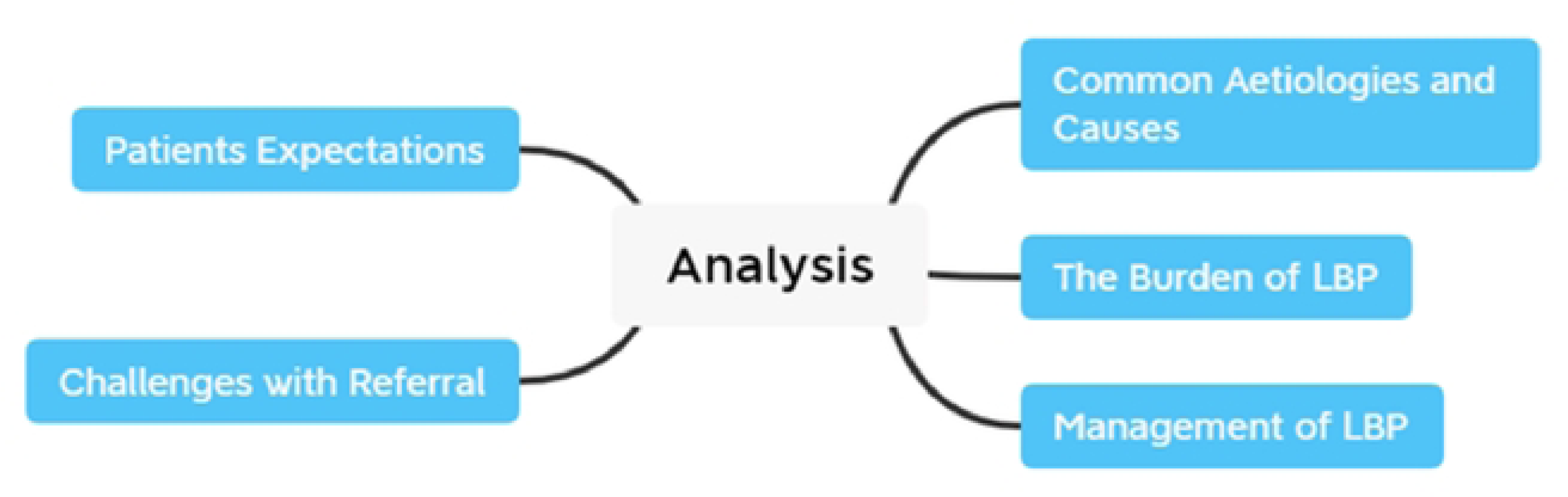
Themes used for thematic analysis.

## The burden of LBP

Low-Back-Pain (LBP) is a common condition among patients attending health centres in rural Uganda, although not always as the primary reason for attending or coexisting with other disorders:

> *“It’s a big problem in my setting because on a daily [basis] if you are attending to 60 clients, about 15 of them will present with this problem”*
>
> *“Many patients, about 70% of our patients, come complaining of other symptoms. Of course in most cases they bring in the back ache but not as a prime symptom”.*

For instance, LBP was a common symptom among women presenting with Pelvic Inflammatory Diseases (PIDs):

> *“ So most of them women of the reproductive age have those afflictions [PID]. So they present with back pain.”*

LBP has an important gender dimension; some participants reported that women are over-represented among patients consulting for LBP, due to coexisting gynaecological disorders or perceived differences in health seeking behaviour, while LBP was seen as “*common in men because of their method of work.”*

> *‘Low back pain is a serious problem, especially in the women compared to men’*
>
> *‘And it’s more common in women than men. I don’t know whether because men don’t seek healthcare so often’*

Chronic LBP was perceived as a common complaint among older people, with patients attending frequently and repeatedly for care:

> *“The elders come back every month complaining of their back.”*
>
> *“In my experience the patients we see, you find that they are too old. You get one old mukade (old woman) comes with low back pain. She will tell you medical documents to show the history. She will tell you I have tried this and that and nothing has helped. You see that she is still in pain.”*

Back pain was perceived by HCWs to be a ‘serious problem’, in terms of its high prevalence, health seeking behaviour, potentially severe underlying causes and severity of presentations.

> *“Backache is a serious problem. Every day I meet such cases whether at my place of work. Even outside this room when I meet someone and they learn, oh, you’re a ‘musawo’ (health worker), I have this problem”*

## Common aetiologies and causes

Several causes or risk/contributing factors for LBP were proposed including age, trauma, disease, obstetric and gynaecological issues, sedentary lifestyle, occupation or farming related factors and lastly, patient factors.

**Working conditions and occupational hazards** are perceived as a major source of LBP. The impact of excessive manual labour and resultant mechanical stress on the spine affects workers, especially subsistence farmers:

> *“They bend too many times”*
>
> *“Also maybe we can add people who carry heavy load. The heavy lifters it could also affect them”*

Female work roles within the household and for income generation were perceived to contribute to low back pain:

> *“Most common group who get lower back pain are women because of the nature of their work, especially being a peasant, at the same time, plus the housework. You find most of the time the woman is the like the soul of the family to to prepare or, to dig for the home. So most of the time they are the ones who come complaining about back pain, maybe like 3 out of 7 I would say are women. And they are more severe, especially when they have then the season maybe of planting or just, normally that’s when they have such a complaint”*

**Age and sedentary time** intersected as contributing factors to LBP, such as office or healthcare workers with sedentary occupations or elders with low levels of physical activity:

> *“Physical inactivity. People who sit a long time, especially the elders, that can dispose them to LBP”.*
>
> *“Also posture. Also some people like as you see more than a hundred patients a day and we see ourselves sitting since morning until lunch time. That one also affects your back”*

**Injuries** as a result of falls or violence were deemed a source of low back pain:

> *“Then I also think of other cause of low back pain: Injuries. Injuries to the spine after accident, sliding”*
>
> *“There are a number of causes; infection, trauma, accidents. Some have been assaulted and beaten, some chronic diseases like diabetes, TB, then PIDs [pelvic inflammatory disorders]-they all cause back pain.”*

As quoted above, participants reported a wide range of **non-musculoskeletal conditions** which manifest as LBP presentations, including serious pathologies such as extra-pulmonary tuberculosis or cancers, and other common benign pathologies, such as pelvic inflammatory diseases.

> *“People they come and they and have PIDs [pelvic inflammatory disorders], inflammatory infections, TB infection, TB of the spine, malignancies, cancers’*

Pelvic inflammatory disorders were perceived to be a common contributor to low back pain as well as other **female specific health conditions** such as pregnancy related low back pain and post caesarean section.

> *“To add on my what my colleagues are saying, about 10 mothers who deliver by caesarian section, most of them come with low back ache. And now most of them are saying maybe they should give us general anaesthesia instead and it is true and it is very common. We as health workers do not know what causes that. After like two years of delivering by caesarian they come back with low back pain.*
>
> *Of course we give them analgesia like Indocid, Diclofenac injection but it does not get cured completely. It keeps coming back and they keep complaining. Yeah”*

**Patient factors** such as poor knowledge and awareness were considered to be risk factors for LBP:

> *“Also think that people lack information on low back pain, how to prevent these thing from happening. In the end they end up doing the wrong thing because they do not have information about what is back pain, how, how it is prevented”*

Patients were reported to have **strong beliefs** about the underlying cause of their LBP, some of which echoed that of health workers in terms of work, age and pregnancy-related low back pain:

> *“There is another one that the community relate back pain to old age because whenever a person complain of back pain they will just say “ hey you’re grand old” so some things that come automatically”*
>
> *“And maybe some of them think the nature of work, for them they’re heavy lifting every day they think you have to have back problem.”*

Participants reported that female patients attributed their back pain to labour and childbirth or menopause:

> *“Surgeries especially for women who have had caesarean section. Many attribute the back ache to the spinal injection.”*
>
> *“Others will say you see because of my age I no longer menstruate and because of that it is disturbing my back. They think it is menopause that is the problem. They may go as far as thinking that maybe if they could bleed, they may get relief in the back”*

Sexual intercourse was also perceived among patients to contribute to back pain

> *“There is also another misconception in the community especially men when they take long without having sexual intercourse, it can also cause low back pain”*
>
> *“Some attribute it to family planning in the case of women, others attribute it to old age, hard work, others attribute it to; especially among the couples, people attribute it to a lack of sexual exercises, and women may say I am too young but my husband is too busy and is not working on me. Backache is a serious problem.”*

## Management of LBP

There is a significant variability in the management practices among clinicians, with an overarching and predominant focus on pain management strategies and onward referral for investigations, usually X-Ray.

Pain management strategies varied among participants and included combinations of medicinal and non-medicinal strategies. The medicinal options encompassed common drugs and some medicines not usually indicated in LBP management:

> *“Ibuprofen, paracetamol, diclofenac,…… Sometimes you can add the vitamins, the B complex. And also sometimes muscle relaxants.”*
>
> *“Managing pain you have to take the history of the patient. So you have to know the type of work the patients does. Is it a man? Is it in the middle age? Depending on the work you may advise to reduce the work and will give some painkillers”*

Non medicinal pain management strategies included general advice to reduce work and recommendations for warm compresses or massage, although participants perceiving these interventions to be ineffective:

> *“Then for the elderly especially we normally encourage them to do warm compress but they say now at home no one is there, my husband is too busy and not around and is no longer interested”*
>
> *“Others have been advised to go for massage, others have been advised to exercise, and still they would come with some little progress.”*

Exercise as part of LBP management was rarely mentioned, with a clear need for further education and training for clinicians in terms of prescription of appropriate exercise and advice relating to physical activity and occupation.

> *“To add to what he has said on referral, when you tell a patient to do exercises, somehow, they be thinking; I am digging and working and so they have the idea that exercises that make you sweat are the actual exercises. I think it would be good for us to gain knowledge on what exact exercises we can teach our patients to relieve their back pain.”*

## Patients beliefs and expectations

Patients’ expectations and beliefs strongly influenced how LBP is managed in HCs, with some patients having strong preferences for specific treatments, particularly medicines, to eradicate their pain:

> *“Most patients actually know the effects of pain killers. For some of our patients, they have taken so many pain killers and when you try to tell them that we should not use drugs, they lose hope and say: “Hati emibazi yona obu nagikozesa ekahwaho (since I have used all the drugs and exhausted them), it means I am going to die”. So they reach an extent of thinking that now the back pain is going to end their life”*
>
> *“Sometimes when they have experienced pain for a long time, they also have relatives who have suffered from back pain and they share advice. So they ask for injections and also they ask for calcium tablets because they associate it with strengthening bones”*

Patients, desperate for pain relief, may self-medicate, procuring medicines from outside of the health system:

> *“It (low back pain) is common and it is a burden. Most of these people which come have had some self medication. In fact, we used to have injections from Tanzania they call Dismas. In the old days Dismas was a very good analgesic given intramuscularly. But in most cases, they go for that.”*

Indeed, LBP is rarely perceived as a complex condition by patients and they have strong expectations that their pain would be quickly resolved:

> *“People expect that when they come you treat them and pain goes as fast as possible. When you tell them it is chronic they don’t understand, they think they should be treated”.*

Advice and exercises alone for LBP was perceived to be unacceptable to patients who may assume that nothing has been done for them and are desperate for fast pain relief to return to manual labour:

> *“When you give them advice they will assume you have not helped enough.”*
>
> *“Patients expect to get drugs. Even if you speak and speak, explain everything and give advice, they feel you have done nothing if they don’t go home with drugs before they can leave the facility.”*
>
> *“Most of them, they are expecting pain relievers. especially someone who’s doing work digging. So when you tell her back needs exercise she would think the worst thing..So she wants. She’s ever taking painkillers, so now she has come to get [them].”*

## Referral Challenges

Onward referral for investigations, usually X-rays, was a dominant theme, in terms of features of patient management and challenges associated with referral pathways. Some clinicians advise patients to pursue x-ray imaging as a first line response, depending on household financial capacity and intergenerational solidarity; for instance “those especially whose grandchildren can afford”.

> *“So when the patients tells you about back pain you good deep, and ask about maybe, as we had seen if this is maybe an infection? Or fracture of structure or any other cause, then, when you rule out some of the causes, then now you can, if you can now it’s really sent for X-ray”*

There is an important loss to follow-up when patients are referred to upper-level facilities, as described by a participant:

“Like 10, you may send like 3, and probably one would manage to go because of the referral system for the x ray”.

An important challenge is the lack of financial resources associated with travel from rural to urban area where referral services are located:

> *“Patients usually go back to the community and they don’t continue on services for reasons such as financing and all that. And then after a certain period you see them coming back to you and they want you to help them.”*

When seeking care at Mbarara Regional Referral Hospital (MRRH), expected long waiting times onsite constitutes a significant barrier. Clinicians reported stories whereby some people “*would go there and see that they sit for good hours and if you’re not patient you’d go back home*”. This repeated phenomenon acts as a deterrent to seek care and reveals the need for key workers to accept referrals. A participant illustrated the problem:

> *“So now the problem is the referral system, because if I’m a health worker, and I referred this patient if possible, if I’m able to connect to someone I’m referring to. This patient gives confidence that when she goes there she’ll be worked on because most of them fear coming here the whole day, she sits and she don’t receive treatment and at the end of the day she was back.”*

Physiotherapy services are found exclusively in Mbarara, and the low numbers of physiotherapists means that it is difficult and expensive to access and in scarce quantities. Accessing these services are costly for patients who may need to travel several times to attend multiple rehabilitation sessions. These constraints paired with the uncertainty of being attended to creates dissatisfaction and refusal to seek care. Older people are particularly at risk: “*They are in the old age and they don’t have things like finance and transport to support them*”.

Because physiotherapy services are underdeveloped, both clinicians and patients are often unaware of the existence and/or the scope of what these services have to offer for LBP, thus nullifying the appropriate referral possibilities:

“Yes, maybe I would say sometimes I’ve been referring these patients, but not specifically to the physios sometimes we say for physician to review for the surgeon”.

Several participants disregard what rehabilitation services could offer to patients with LBP. The following quote illustrates how LBP is commonly approached only with a biomedical lens:

> *“personally, I never regarded physio as a way of managing LBP. I Also use it to look at a line dimension X ray and maybe surgery. But now it’s good that we know physio can be a treatment”.*

One participant advocated for a decentralisation of rehabilitation services to the communities to overcome the multiple referral challenges:

> *“To me all these services should be expanded to all the HCs because it’s where we get these patients, most of them don’t know that they can benefit from physiotherapy”.*

## Discussion

This study sought to understand the experiences, perceptions and practices of frontline health care workers in rural health centres in Uganda in relation to LBP management. Findings demonstrate that LBP is a common and persistent complaint among patients attending rural health centres, with severe impacts on lives and livelihoods. Despite this, LBP and musculoskeletal disorders are not a priority for health service delivery and planning, despite a renewed focus on non-communicable disease and injuries in the Ministry of Health National Health Strategy [22].

Physical work was deemed to be a root cause of LBP, and in turn, LBP had devastating impacts on people’s livelihoods, with women severely impacted. The need for clinicians to provide ‘quick’ working solutions was deemed critical to enable people to return to work and social roles. There was a perception that evidence based strategies such as exercise and advice may not be tolerated or even appropriate for an already highly active population. Evidence based LBP management is derived from research from HICs, and therefore tuned to populations who are in the main sedentary-This is in stark contrast to people living in rural Uganda, where, as evidenced in our study and others [23] people living in rural communities are highly active due to the manual nature of their occupations. There is therefore an urgent need to develop and adapt evidence based interventions for LBP that are appropriate to the population and context in rural Uganda and crucially, deep consideration of how these interventions can be implemented, mainstreamed and delivered in rural health centres within the resources and time available, to be effective.

Currently, front-line clinicians are ill-equipped to manage LBP in Uganda. Readily accessible management options are limited to basic drugs and investigations such as Xray and blood tests. These strategies are free to patients in the public hospital system but are subject to long waiting times. Furthermore, these strategies (over reliance on medication and referral for X-ray) are at odds with evidence based practice for non-specific LBP.

Essentially, the management of LBP can be summarised as mainly transactional and passive (i.e. treatment is limited to the consultation interaction, with little capacity to empower patients to self manage), highlighting a dominant medical model of LBP care, strongly contrasting with evidence based biopsychosocial approaches [15]. Strong dependence on medication was evident both from how participants report their management strategies and their perceptions of patient expectations. In the absence of available and affordable alternatives, such as education and rehabilitation, health workers tend to prescribe a large amount and diversity of medication, as observed in other LMIC contexts [24]. A study investigating antibiotics use in rural eastern Uganda found that for patients presenting with back pain, “antibiotics were often prescribed in addition to painkillers to treat possible infections” [25]. The routine prescription of antibiotics for pain was perceived to enable return to physical work, despite health workers and patients’ concern about harmful side-effects. This study also found that local residents frequently stock-piled medicines at home, including previously prescribed antibiotics, which were commonly used to self-manage pain. The researchers discuss how prescription, delivery, storage and use of medications such as painkillers are intricately linked to the socio-economic constraints within which people live and their expectations when seeking care, [25, 26].

Patients’ dependence on painkillers was evident in our findings from both their reported efforts to secure medications, even from outside of the country and their dissatisfaction if medication was not prescribed. Painkillers usage is frequent in Uganda and opioids drugs “such as morphine, tramadol and pethidine, are easily available to the Ugandan public.” [27]. This availability accentuates the “growing global problem of controlled prescription drugs use disorders” which is aggravated by “.low compliance to regulations on CPDs among private pharmacies in Uganda” [28]. Despite the widespread availability and misuse of opioids such as tramadol, the policy context and oversight has been described as incoherent, thus creating “a tautological problem in education” amongst healthcare professionals [27], further worsening the problem of inappropriate prescribing.

X-ray referral was a common diagnosis strategy, although at odds with EBP guidelines, whereby unnecessary radiology is deemed harmful and may contribute to increasing fear avoidance patterns and pain related psychological distress, further increasing the risk of persistent pain and LBP related disability [29]. Furthermore, unnecessary referral to Regional Referral Hospital for x-ray places a considerable financial burden on patients in terms of direct out of pocket health care expenses and travel costs and on the health system, in terms of wasting valuable radiology resources.

Even when referral to specialist services for further investigation is essential (e.g. suspected underlying serious pathology) or treatment is appropriate, clinicians highlighted the access challenges for patients. MRI or CT investigation is extremely costly in Uganda (e.g. approximately 7-12% of the average annual income) and constitutes an out of pocket expense for patients [30]. This lack of financial capacity coupled with transport and accommodation expenses and the difficulty for patients to navigate the healthcare system when referred were pointed out as key challenges. Lack of rehabilitation workers paired with flawed health referral information systems constituted two structural challenges for efficient management when patients reached the referral hospital. These results reflect findings from Sharma and colleagues, investigating LBP management practices in 32 LMICs [24].

The findings from this study underline the urgent need for enhanced education and training for frontline clinicians to manage LBP through enhanced and appropriate prescribing and adoption of basic rehabilitation skills.

Education and training of health professionals for pain management at all health system levels is crucial to ensure appropriate and safe prescribing. Training should include therapeutic counselling with patients with persistent pain to empower patients to manage pain and prevent medication abuse. However, any efforts focusing solely on healthcare professionals will have very limited impact if it does not take into account the perspectives and motivations of prescribers and patients and the socio-economic context within which these practices are embedded. Rigorous investigation is required to understand and address the structural components influencing pain medicine prescription and consumption to avoid falling into an individualist behaviour change approach to a profoundly social issue [31].

Evidence-based and context-specific rehabilitation interventions need to be developed and adapted so that they can be delivered within the time and resource constraints of the health workforce and to ensure that rehabilitation interventions are relevant and acceptable to patients in the context of rural Uganda. Such interventions need to be co-designed and tested in this context, to ensure that they are feasible for uptake and implementation but also tailored to meet people ’s needs so that they are effective. In the long term, the rehabilitation workforce should be enlarged and rehabilitation services mainstreamed into primary health care at the community level.

To our knowledge, this is the first study to focus on LBP management in rural Uganda. It showed that determinants of LBP management and outcomes act at the micro-level and the macro-level. At the micro-level, the clinicians’ knowledge and the services available at a given facility acted as modulating factors of LBP management. At the macro-level, the healthcare system’s complexity, lack of transport and the high costs associated with healthcare access paired with health workforce shortages and poor health information system were identified as major determinants of the care pathway.

The restricted geographical diversity of this study creates a limit to our ability to generalise the understanding of the contextual determinants. The fact that interviewers were physiotherapists may have biased participants who may have felt obliged to mention rehabilitation more than they normally would. Furthermore, a workshop conducted to introduce the broader BACKTRACK project prior to the FGDs may have influenced responses about LBP management.

This study provides a strong understanding of the landscape and scale of the LBP problem in rural Uganda. It has shown the challenges of managing LBP at the intra-facility level and the several inter-facility referral challenges patients experienced. It can inform innovative solutions aiming to address the burden of LBP in rural areas. Findings from this study and a concurrent study of LBP patients have informed the BACKTRACK project in the co-design, development and evaluation of a mHealth clinical decision support system for LBP. Future research aims to co-create an evidence-based e-learning programme for HCWs, tuned to the Ugandan context, to enhance knowledge and skills to manage LBP in the rural setting and develop an implementation plan to support adoption and uptake of the programme.

As the research team was predominantly physiotherapy researchers, at the outset the focus of the study was related to rehabilitation. Crucially, this study findings have revealed the necessity to investigate comprehensively how painkillers are supplied, prescribed, delivered and used by clinicians and patients. Further investigation is required to understand the perspectives of users, pharmacists, doctors as well as policy-makers and regulators. Finally, there is a need to investigate the perspectives of doctors at referral points about the use of non-pharmacological means to manage back pain, such as physical rehabilitation, as well as the relative effectiveness of painkillers in the management of chronic low back pain in the long term.

## Data Availability

Data are not publicly available because participants did not give explicit consent for the data to be held in a public repository

## Acknowledgements

The research team would like to express their utmost gratitude to all clinicians involved in the projects for their time and efforts.

